# Methods to engage ‘under-served’ people with chronic disease in contributing to research: A scoping review and stakeholder prioritisation

**DOI:** 10.1101/2025.03.06.25323512

**Authors:** Paula Boeira, Tamar Avades, Amanda Clements, Ashwin Dhanda

## Abstract

**Background:** People with lived experience of chronic illness have much to contribute to all stages of research from design to dissemination. However, not everyone is able to engage in research, with low representation from under-served groups. People from these groups use their lived experience to provide a different perspective compared to people that traditionally engage with research. The aim of this scoping review is to identify and evaluate the current evidence on engaging under-served populations with chronic disease in research.

**Methods:** A scoping review was conducted in accordance with PRISMAScR guidance. Two databases of peer-reviewed literature (Scopus and Web of Science), grey literature and websites were searched from 2009 to present. We included any study type that used at least one intervention to engage people with chronic illness from an under-served group in research.

**Results:** Five studies reporting ten methods of engagement were included. The most frequently described method (n=4) was ensuring accessibility to research studies (e.g. location, transportation, timing). Three studies tested the provision of financial incentives. Other methods described included ensuring cultural sensitivity and using peers to support participants. The methods of engagement identified in this scoping review were discussed by stakeholders from relevant community organisations and patient representatives, which informed the interpretation of data.

**Conclusions:** Few methods of engagement of under-served people in research have been tested. Many factors contribute to the underrepresentation of minorities in clinical research. Constant assessment to improve efficiency in identifying, enrolling, and engaging minorities in clinical research is important.

## INTRODUCTION

Public and patient engagement is an essential part of the research process and should be embedded in all aspects of a research study from developing the research question to dissemination of findings (National Institute for Health and Care Research Report, 2015). However, the people and their social network who are affected by a research study should all have an opportunity to engage with it. The challenge for researchers is to enable all stakeholders to have a voice and not only rely on self-selected people who may not be representative of the population being studied.

The under-served population is specific to the research project. It is a broad term that could include people experiencing deprivation or homelessness, sex workers, low literacy groups, people with disabilities, people with substance or alcohol dependence, ethnic minorities, people in the social justice system and people in certain locations (National Institute for Health and Care Research Report, 2021). Other terms such as ‘hard-to-reach’, under-represented or vulnerable have been used to describe these populations (Shaghaghi, Bhopal & Sheikh, 2011). However, these descriptors imply that the problem in engagement is due to the population itself rather than the methods of engagement used.

There are numerous reasons why under-served groups do not engage in research. Many people in these groups do not have access to transportation, do not have means to be contacted such as a mobile phone, fear being stigmatised and discriminated against or have low health literacy (Ellis, 2021). Understanding these challenges is essential for developing strategies to increase engagement for under-served groups.

Studies have been conducted to test the effectiveness of interventions to improve participant engagement in trials. A Cochrane systematic review has summarised methods to enhance retention in randomised clinical trials finding only a few methods that have been tested with moderate-certainty evidence (Gillies et al,2021). These included financial incentives with clinic reminders, self-sampling kits and giving a pen at recruitment. However, this review focused only on randomised trials and did not include under-served populations specifically.

As researchers in the field of liver disease, we frequently encounter challenges in engaging under-served groups of people in research including people with alcohol and drug dependence, ethnic minorities, people experiencing homelessness and those from deprived areas with a high prevalence of liver disease. We wanted to understand how to engage such people in contributing to research. However, an initial search restricted to liver disease revealed no relevant literature. Therefore, we broadened the review to include people with any form of chronic disease.

More than 15 million people in England suffer with chronic illnesses (Stafford et al, 2018). There is health inequality in the prevalence of long-term conditions, which is 60% higher in people living in socioeconomically deprived areas than non-deprived areas (Department of Health Report, 2012). Chronic diseases have an earlier onset in people living in deprivation and are more common in ethnic minorities (National Institute for Health and Care Research Report, 2021). This scoping review explores what is known about engaging under-served populations with chronic disease in research.

## METHODS

The scoping review was conducted in accordance with the Joanne Briggs Institute methodology for scoping reviews (Peters et al, 2020). A full protocol was developed and published on the Open Science Framework (https://osf.io/mu26c/).

The scoping review sought to answer the question: “What methods of research engagement have been tested in people with chronic illness from under-served communities?” We considered all studies evaluating an intervention targeting research engagement including randomised controlled trials, non-randomised trials, before and after studies, interrupted time-series studies analytical observational studies.

We performed searches, from 2009 to present, in two databases of peer-reviewed literature, Web of Science and Scopus. Web of Science includes the Medline database and covers similar literature to EMBASE. We applied the same search strings to identify grey literature reported on 27 pre-defined websites of UK government, health service or charity organisations (see Supplementary Methods for full list). The following search blocks were combined:

### 1. Population of interest

Disadvantaged, deprived, homeless, drug users, alcoholic, alcohol dependent, ethnic*, hard to reach, under-represented, substance misusers (users), criminal offenders, prostitutes, injection drug users, men who have sex with men, sex workers, migrants, prisoners, people living with HIV, asylum seekers, refugees, vulnerable, hidden, difficult to reach, poor, low income, gay, lesbian, bisexual and transgender, low literacy group, Mental illness, People with a disability

### 2. Engagement with research (terms for Engagement AND Research were combined)

#### ENGAGEMENT

Engage*, participant*, design*, co-design, co-production*, PPI, PPIE, public involvement

#### RESEARCH

Research, trial, study, intervention, health research, clinical, medical, recruitment, survey, sampling, retention, data collection, analysis

### 3. Chronic disease (terms for Chronic AND Disease were combined)

#### CHRONIC

Chronic*, long-term, long-lasting, persistent DISEASE

Disease, ill*, condition, disorder, health problem, medical condition, sickness

### Data analysis and interpretation

Data were categorised using an analytical framework that was agreed between the research team. A summary of results were presented to stakeholders in the management of people with chronic liver disease including representatives from homelessness services, alcohol services and voluntary and social enterprises (supplementary materials). Discussion with the stakeholder group informed the interpretation of data and consensus was reached regarding the prioritisation of methods to apply to people with chronic liver disease.

## RESULTS

The Scopus search returned 1123 articles, from which five duplicates were removed. The Web of Science search resulted in 770 articles. Titles for the remaining articles were evaluated for relevance. Subsequently, fifty-five papers were found to meet the inclusion criteria and abstracts were assessed to determine their eligibility. Thirty-six were excluded after abstract screening. On full review of papers, 14 were excluded as they focused on medication adherence rather than engagement with research or did not include patients with chronic diseases and therefore were not relevant for this review. Finally, five papers were eligible for inclusion in the scoping review and read in full (Figure 1).

**Figure 1.**
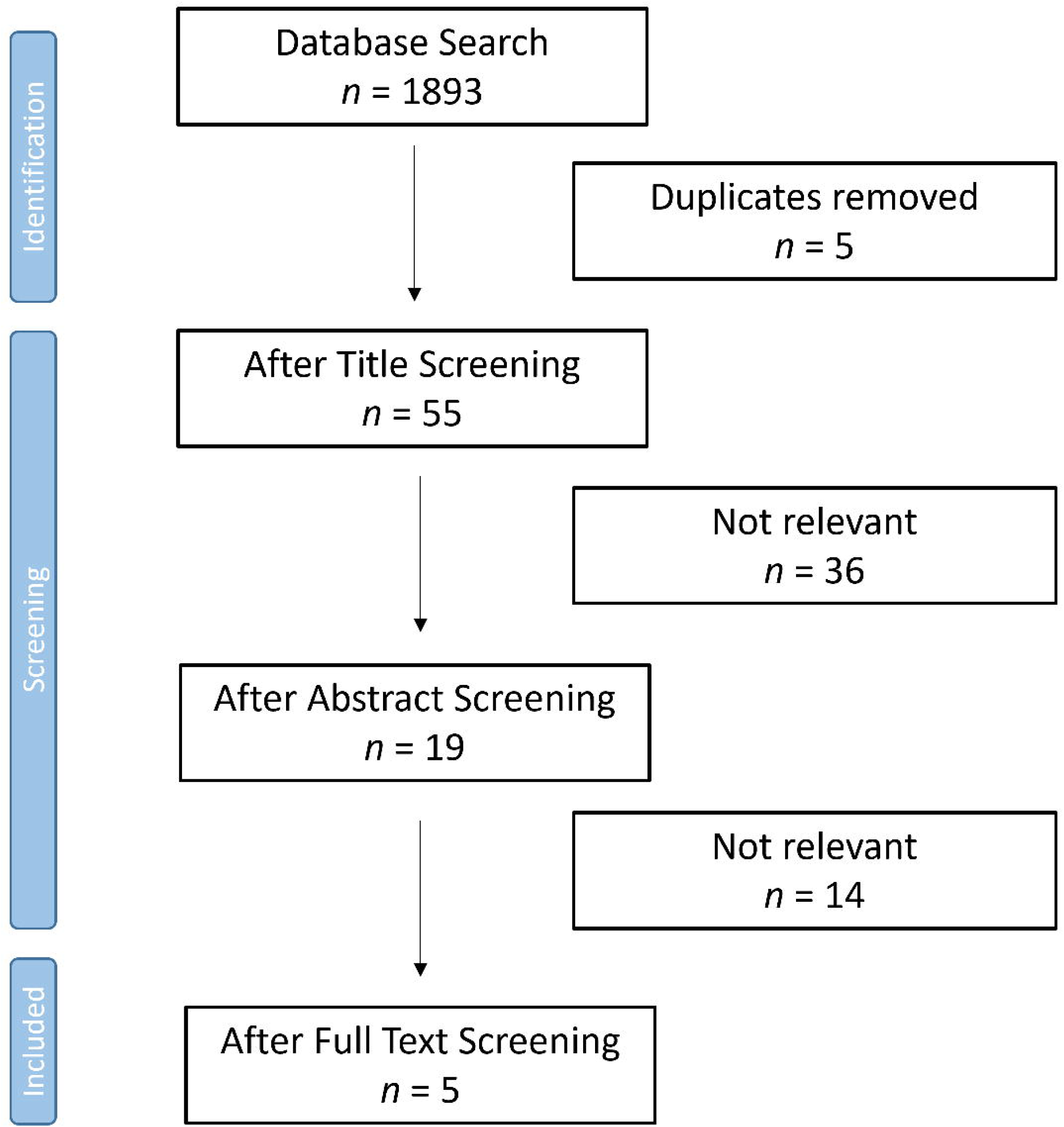
PRISMA flow chart for database searches. PRISMA, Preferred Reporting Items for Systematic Reviews and Meta-Analyses.

From 27 websites, we identified 20 online reports with engagement methods. After review of the full reports, no website met our inclusion criteria (Figure 2).

**Figure 2.**
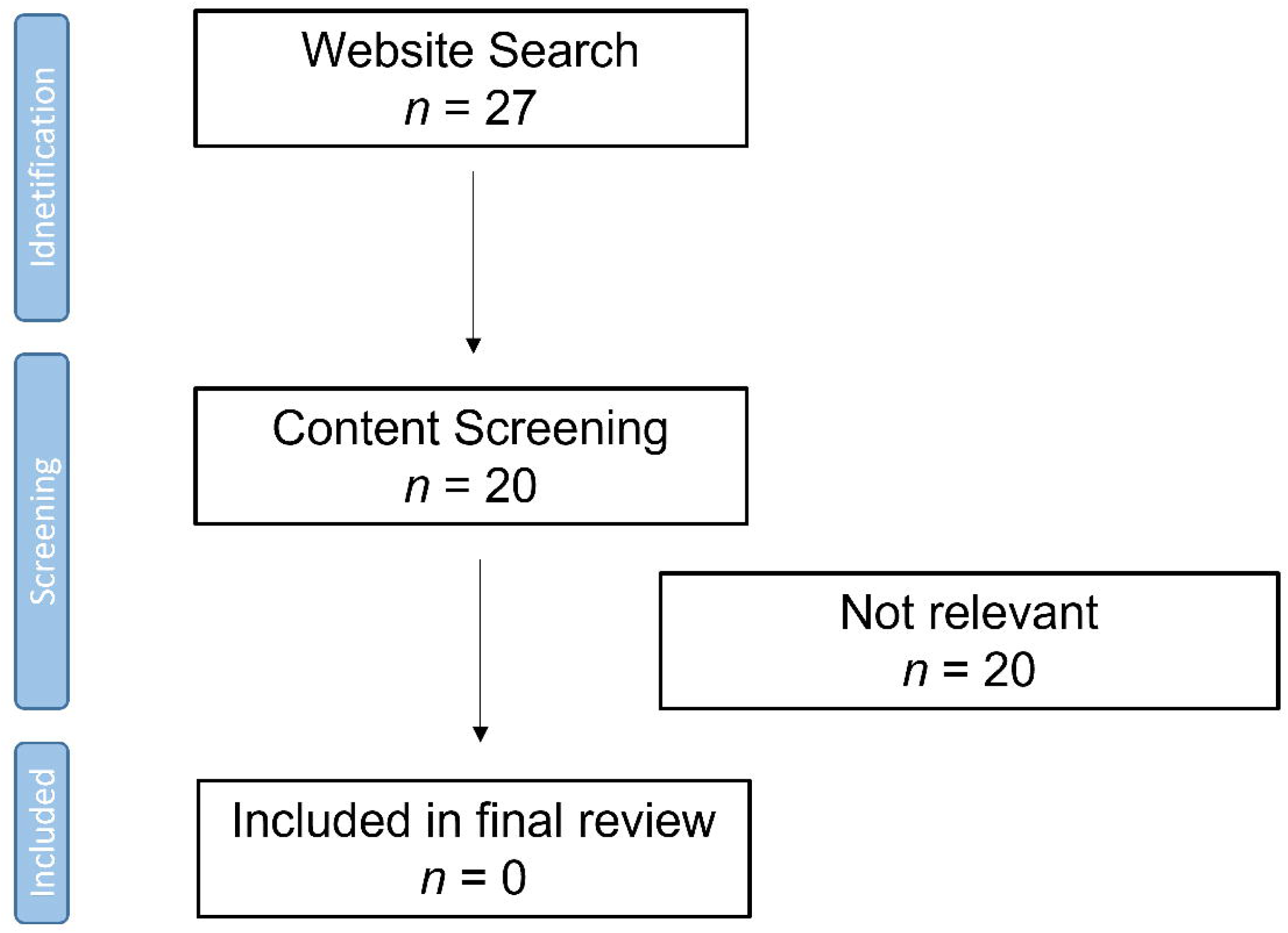
PRISMA flow chart for charity/government website searches. PRISMA, Preferred Reporting Items for Systematic Reviews and Meta-Analyses.

### Study characteristics

Of the five eligible articles identified from database searches, four were conducted in the United States and one in Ireland. Concerning study design, four studies were interventional and prospective, and one was observational and retrospective.

### Targeted group

The most common underrepresented population included in three studies was ethnic groups and older adults were assessed by two. Regarding chronic disease, of the five articles, two studies focused on multiple chronic conditions, one study included type 2 diabetes, one on stroke and one on systemic lupus erythematosis.

### Categorisation of engagement methods

Using the analytic framework we grouped methods of engagement into three main themes: accessibility, communication and relationships, and incentives (Table 1). Within these themes tens methods of engagement were described: convenient location, transportation support, flexible hours, cultural adaptations, frequent contact with healthcare providers, health focused peer community approach, reminders, a user centred design, financial compensation and non-monetary gifts (Table 1).

**Table 1.** Methods of engagement identified in scoping review. Methods are grouped into three themes using an analytical framework.

|  | Methods of engagement | Papers | Counts |
| --- | --- | --- | --- |
| Accessibility | Convenient location | Doyle<br>Drenkard<br>Magwood<br>Sefcik | 4 |
|  | Transportation support | Drenkard | 1 |
|  | Flexible hours | Drenkard | 1 |
| Relationships and communication | Cultural adaptations | Osborn<br>Drenkard | 2 |
|  | Frequent contact with healthcare providers | Sefcik<br>Doyle | 2 |
|  | Health focused peer community approach | Sefcik | 1 |
|  | Reminders | Drenkard<br>Doyle | 2 |
|  | User centred design | Doyle | 1 |
| Incentives | Financial compensation | Osborn<br>Drenkard<br>Sefcik | 3 |
|  | Non-monetary gifts | Drenkard | 1 |

The commonest recommended method was the accessibility of the research, with four papers describing the provisions made to conduct research in locations that were convenient and familiar to participants. Accessibility was further enhanced with transportation being supplied in one study and flexible hours being offered to attend workshops. These methods minimise the burden to the participants.

Second was the importance of a strong relationship with a trusted research team that was knowledgeable on cultural and social issues, and who provided regular contact with the participants. Reminders by phone, text message or email were recommended to encourage attendance at research meetings, but this applies more to people already participating in research. Cultural adaptions and understanding of social norms of that particular community was beneficial in African-American women and Puerto-Rican patients with diabetes. Elderly patients with chronic conditions favoured a peer-led approach in the community. Studies suggested that careful consideration is given to who and when to approach people and how to do it in a culturally and socially sensitive manner.

Third was the use of monetary incentives, which was reported in three studies, with two studies offering gift cards of increasing value at different stages of completion.

#### Accessibility

Delivering an intervention at a convenient time may overcome barriers such as job schedules and childcare needs. A non-significant higher completion rate was observed for workshops delivered on Saturdays (76.6%) compared with weekdays (74.0%); afternoon and Saturday workshops enabled attendance of those who had medical appointments, full-time jobs or caregiver commitments (Drenkard et al, 2020).

A convenient location for the intervention facilitated engagement: a study performed by researchers who visited the participant’s home facilitated engagement (Doyle et al, 2021). Interventions were delivered in the most convenient locations to the participants including libraries, health centres, churches, community centres nearby study participants’ residency, and healthcare facilities that were familiar to the participants (Drenkard et al, 2020).

Sefcik et al (2018) also reported that an accessible location was chosen to facilitate participants’ visits, in addition the study explains that in-home nurse visitation was highly effective (Sefcik et al, 2018). Likewise, Magwood et al (2021) described that interview groups were held in various locations across the 4 neighbouring counties to facilitate access from rural areas and to encourage participation from those who preferred not to travel to the academic medical centre (Magwood et al, 2021).

To overcome the challenges faced with poor recruitment in the Chronic Disease Self-Management Program, the participants were offered flexible transportation support (Drenkard et al, 2020). Together with a non-monetary gift and reminders, retention increased from 64.7% to 80.0%. The research coordinators also provided flexibility by offering alternative workshops to improve their retention in the trial.

#### Incentives

Three studies reported that gift cards were provided to cover participation and transportation costs (Drenkard et al, 2020; Osborn et al, 2010; Sefcik et al, 2018). Compensation ranged from $20 in total to $50 per visit (Drenkard et al, 2020; Osborn et al, 2010). One study also sent participants a study bag and greeting card to encourage retention (Drenkard et al, 2020).

#### Reminders

SMS text messages, emails, calendar alerts, and voice messages can help patients stay on track with the study protocol by improving patient adherence, compliance, and retention. As part of a wider retention strategy, sending reminders by emails text message or phone to participants increased the completion rate of workshops (Drenkard et al, 2020). Monthly phone calls were used to keep participants engaged in a separate study (Doyle et al, 2021).

It was recognised that participants missed study workshops due to forgetfulness. Weekly phone calls provided a friendly reminder of their workshops (Drenkard et al, 2020).

#### Cultural adaptations

Two studies discussed cultural adaptations as a method of engagement in Puerto Ricans and African-American women, respectively (Drenkard et al, 2020; Osborn et al, 2010). Osborn et al (2010) described applying the Information-Motivation-Behavioural skills (IMB) model on Puerto Ricans with type two diabetes. Those that were recruited into the intervention arm received sessions that were delivered by a bilingual healthcare professional. In the information session, participants were given information on which culturally relevant foods increase their blood sugar. In the final session, participants were encouraged to reflect on how their cultural situation (e.g., cultural norms and expectations) impacted on their behaviour towards their health. Only those in the intervention arm had a significant decrease in their glycaemic control (Osborn et al, 2010).

Drenkard et al (2020) identified that a barrier to patients being lost to follow up following recruitment or failing to show up was secondary to a misalignment of the objectives between the participants and the intervention program (Drenkard et al, 2020). They developed cultural-relevant materials for the participants to help them understand the objectives of the study. The materials included information regarding the recruitment process and short videos to highlight the main messages. They did not describe what constituted ‘culturally-relevant’ material, but their aim was to ensure the objectives of the patients and the program were aligned (Drenkard et al, 2020).

#### Frequent contact with healthcare providers

Sefcik et al (2018) conducted a qualitative study to evaluate patient experiences of the Health Quality Partners (HQP) organisation, which offered a dedicated nurse to reduce hospitalisations (Sefcik et al, 2018). They also offered a health-focused peer approach in the community (see below). Over a twelve-year period, a variety of trials were conducted to measure the efficacy of coordinated care interventions on reducing hospital admissions in patients aged 65 and over, with a chronic condition. The patients in the HQP intervention arm had a dedicated nurse assigned to them who was responsible for coordinating their care (Sefcik et al, 2018). The initial study found that those who had face-to-face contact with a dedicated nurse were more likely to remain enrolled in the trial due to provision of both health related and social support (Toles et al, 2017).

Frequent contact with a healthcare provider was also noted to be a potential facilitator in engagement in the Pro-ACT trial (Doyle et al, 2021). A digital platform enabled elderly patients to manage their multiple co-morbidities by entering data that was remotely monitored, providing a regular connection to the research team (Doyle et al, 2021).

#### Health focused peer community approach

The HQP program described by Sefcik et al (2018) was developed to engage elderly people with multiple chronic conditions (Sefcik et al, 2018). As well as providing a dedicated nurse, participants had access to a health-focused peer community which enabled them to share experiences of their health and develop new skills.

Participants reported a positive impact the community had on their health, with regards to an improvement in their diet as well as physical activity. Participants were able to learn new skills through their peers, but also through materials that were specific to their health needs and conditions. Development of relationships further encouraged their engagement with bettering their lifestyles (Sefcik et al, 2018).

#### User centred design

Doyle et al (2021) reported that the success of their Pro-ACT platform was enhanced by focusing the design on the users, which in this case were elderly patients with multiple chronic conditions. They achieved this by simplifying the data collection onto one platform and providing technical support if needed (Doyle et al, 2021).

Previous research stipulated that unfamiliarity with technology poses a barrier for elderly patients, who would not be accepting of this method (Heart & Kalderon, 2013). The Pro-ACT platform was designed following focus groups and interviews with patients, as well as enabling patients to test the platform to update the design. They concluded that understanding a patient’s needs contributed to the high level of engagement from patients (Heart & Kalderon, 2013).

### Prioritisation of methods to engage people with chronic liver disease

The stakeholder group reviewed the engagement methods discussed. They considered the most important methods to engage people were:

1. Information. The correct information in the correct medium should be provided including consideration of other forms of communication such as cartoons or videos. This needs to be personalised to the individual.
2. Approach. This should be done via a peer or someone in their care network.

The person needs to be authentic and must be in their environment.

The agreed approach to engage under-represented people with chronic liver disease is to adopt a method tailored to the specific group of interest. This should involve identifying champions or peers in existing groups and organisations, training them in the research and asking them to identify and recruit participants to a study using appropriate and accessible information designed specifically for the people of interest.

## DISCUSSION

There is a strong rationale to involve patients in all aspects of research, from identifying the research question to disseminating findings. It can lead to tackling the most relevant research questions, better designed studies, enhanced recruitment and retention, and improved implementation and dissemination of study results (Brett et al, 2014). The National Institute of Health and Social Care Research (NIHR) has recognised this and developed UK national standards for patient involvement in research (National Institute of Health and Social Care Research, 2019). One standard relates to inclusive opportunities and states: ‘Offer public involvement opportunities that are accessible and that reach people and groups according to research needs’. However, methods for achieving this are poorly defined.

Our scoping review has confirmed that there is a lack of studies assessing the effectiveness of methods to engage under-served groups with chronic disease. We identified and analysed five peer-reviewed publications that described how they improved engagement of under-served populations. These studies have revealed some common methods that have been tested. The most frequently described method was to ensure that the study was accessible to all participants with consideration given to location, timing and social factors. Other methods include developing a strong relationship with the community of interest, providing incentives and sending reminders.

Most of the identified interventions have been tested using observational methods and qualitative interviews with participants and researchers. The absence of high-quality evidence for interventions to improve engagement in under-served groups suggests that this is not considered a priority research area or that there are barriers to conducting such studies.

The main barrier is likely to be the specificity of any intervention; it is designed for a specific population for a single study and may not be easily translated to other scenarios. This is highlighted by the findings of this scoping review in which no two interventions were identical. Additionally, some of the methods require an investment in time and people to build meaningful relationships between researchers and under-served groups. These costs need to be accounted for in study design and funding that may be considered barriers for researchers (Domecq et al, 2014).

A scoping review methodology was used to identify the most relevant published studies to this review. Our search strategy included the term ethnic; however, this term is very limiting, and relevant studies may have been missed if they included a specific ethnicity. Our focus was interventions to increase engagement of underserved populations with chronic conditions; the restrictive inclusion criteria led to many studies being excluded.

In conclusion, few studies have been published that test methods to engage under-served populations in research. Those that have, report a range of interventions specific to the research project. Researchers should consider a tailored approach to engagement using one or more of the identified methods and test their effectiveness within the context of the planned study.

## Data availability

The scoping review protocol is available on the Open Science Framework (https://osf.io/mu26c/). All other analysis and data are presented in this manuscript or in the Supplementary Material.

## AUTHOR CONTRIBUTIONS

Ashwin Dhanda conceived the study. Paula Boeira conducted all the searches. Paula Boeira, Tamar Avades and Ashwin Dhanda were involved in data extraction and analysis. All authors contributed to the interpretation of results and drafting the manuscript and approved the final version.

## Notes

**Grant support** This study was supported by a University of Plymouth Explore Engagement Award to Dr Dhanda

### Competing Interest Statement

The authors have declared no competing interest.

### Funding Statement

This study was supported by a University of Plymouth Explore Engagement Award to Dr. Dhanda

